# ZIKA VIRUS VERTICAL TRANSMISSION IN CHILDREN WITH CONFIRMED ANTENATAL EXPOSURE

**DOI:** 10.1101/2020.02.26.20028399

**Authors:** Patrícia Brasil, Zilton Vasconcelos, Tara Kerin, Claudia Raja Gabaglia, Ieda P. Ribeiro, Myrna C. Bonaldo, Luana Damasceno, Marcos V. Pone, Sheila Pone, Andrea Zin, Irena Tsui, Kristina Adachi, Jose Paulo Pereira, Stephanie L. Gaw, Liege Carvalho, Denise C. Cunha, Leticia Guida, Mirza Rocha, James D. Cherry, Lulan Wang, Saba Aliyari, Genhong Cheng, Suan-Sin Foo, Weiqiang Chen, Jae Jung, Elizabeth Brickley, Maria Elisabeth L. Moreira, Karin Nielsen-Saines

## Abstract

**Background:** *In utero* transmission of Zika virus (ZIKV) can lead to adverse infant outcomes, but vertical transmission rates are unknown.

**Methods:** Antenatally ZIKV-exposed children were followed prospectively since the time of the Rio de Janeiro epidemic in 2015-16. Serum and urine specimens were collected from infants from birth throughout the first year of life. Specimens were tested by quantitative reverse transcriptase polymerase chain reaction (PCR) and/or IgM antibody capture Zika MAC-ELISA. Infants had neurodevelopmental evaluations, brain imaging, eye examinations, and hearing assessments.

**Results:** Over time 130 *in utero* ZIKV-exposed (mothers PCR+) children were tested with 407 specimens evaluated: 161 sera were tested by PCR and IgM assays, 85 urines by PCR; 84 children (65%) were positive in at least one assay. Among 94 children tested within 3 months of age, 70% were positive (39% serum PCR, 48% urine PCR, 39% IgM). After 3 months, 33% were positive by any laboratory method. Five children were intermittently PCR+ beyond 200 days of life. Concordance between IgM and PCR results was 52%, sensitivity 65%, specificity 40% (with any positive PCR result as the gold standard); IgM and serum PCR were 61% concordant; serum and urine PCR 55%. Most children (65%) were clinically normal. Positive results were seen in 29 of 45 children (64%) with abnormal findings and 55 of 85 normal children (65%), p=0·98. Earlier maternal trimester of infection was associated with positive infant laboratory results but not infant clinical disease (p=0·04).

**Conclusions:** ZIKV has a high *in utero* transmission rate. Laboratory confirmed infection is not necessarily associated with abnormal infant findings.

---

In 2015, Zika virus (ZIKV) reached Rio de Janeiro, Brazil and local transmission of the virus was quickly confirmed^1^. At the time, our group had an ongoing study evaluating outcomes in pregnant women who developed a rash due to arboviral infections. With the onset of the ZIKV epidemic, we established a longitudinal cohort of pregnant women who presented with a rash within the prior 5 days and were found to be ZIKV positive in blood or urine by quantitative reverse transcriptase polymerase chain reaction (QRT-PCR) at the time of presentation^2^. We followed these women throughout pregnancy to delivery and reported on pregnancy outcomes^3,4^. Subsequently we reported outcomes of their children in the first months and years of life including physical findings, neurologic examinations, neuroimaging results, complete eye exams, hearing assessments and neurodevelopmental outcomes^4–8^.

Although all infants in this prospective Zika cohort were exposed *in utero* to maternal ZIKV infection, it is difficult to ascertain the true rate of ZIKV vertical transmission without infant laboratory results, as the majority of ZIKV exposed children are not born with severe clinical features of congenital Zika syndrome^3^,^9^. Amniocentesis to detect ZIKV by RT-PCR in the amniotic fluid can confirm vertical transmission prenatally although this procedure is uncommonly performed in Brazil^8^. There is limited data on the sensitivity and specificity of testing after birth to confirm vertical transmission. In adults, a confirmed laboratory diagnosis of ZIKV is challenging, as there is a brief window period where virus detection in plasma or urine occurs^10^. Zika serologic testing has been further complicated in endemic areas by the cross-reactivity between ZIKV IgG antibodies and antibodies against dengue virus serotypes; in that scenario most surveillance systems rely on clinical criteria to identify cases^11,12^.

We sought to determine the utility of molecular and serologic testing in the diagnosis of mother-to-child-transmission of ZIKV. We report the frequency of ZIKV PCR and ZIKV-specific IgM detection in the serum and urine of children with confirmed prenatal ZIKV exposure. We also analyzed whether positive infant results for ZIKV were associated with abnormal pediatric outcomes.

## METHODS

### Study Population

The study population was comprised of infants born to women enrolled in a longitudinal cohort of confirmed ZIKV infection during pregnancy^2,3^, for whom postnatal samples of blood and/or urine were collected for ZIKV laboratory diagnosis. Gestational age of infection was estimated based on the day the pregnant woman first presented with the rash due to ZIKV infection confirmed by PCR in blood or urine. All children were followed at the Fundação Oswaldo Cruz (Fiocruz) in Rio de Janeiro and were enrolled from December 2015 to December 2016.

### ZIKV PCR Detection

Infant serum and urine specimens were obtained following parental signed informed consent. Serum was collected by standard phlebotomy procedures and processed immediately for PCR while additional aliquots were stored at – 80C for subsequent testing for Zika IgM. Urine specimens collected by bagged urine collection, spun in a refrigerated centrifuge for 10 minutes, and the supernatant was aliquoted and processed immediately for PCR. Both serum and urine specimens were tested by qRT-PCR amplification assay for ZIKV using the TaqMan probe (Applied Biosystems) for detection and absolute quantification of ZIKV as previously described^2,3,12^. Laboratory testing sites included the research laboratory of the Instituto Fernandes Figueira Hospital (IFF) and the Laboratory of Molecular Biology of Flaviviruses, Fiocruz.

### ZIKV IgM Detection

ZIKV serologic testing of infants was performed in serum aliquots using IgM antibody capture Zika MAC-ELISA from the Centers for Disease Control and Prevention (CDC, Fort Collins, CO, EUA) according to manufacturer instructions^13^.

### Pediatric Outcomes

Pediatric outcomes (normal versus abnormal) were determined based on specific clinical findings.

### Early infant findings

Were defined based on the presence of any of the following medical findings in the first 3 months of life: (1) Microcephaly (MC) defined as head circumference Z-score < -2 (moderate) and < -3 (severe)^14^; (2) Small for gestational age (SGA) at birth based on sex-specific curves by Intergrowth-21^15^; (3) Abnormal eye findings following a complete exam with funduscopic evaluation performed by pediatric ophthalmologists as previously described^16–19^; (4) Abnormal hearing assessments evaluated through brainstem evoked response audiometry (BERA)^3,4,5^; (5) Very abnormal neurological exam, with a constellation of findings on repeated physical examination including a combination of: hyper-or hypotonia, clonus, contractures/arthrogryposis, seizures, continuous irritability (inconsolable crying)^2–7^; Abnormal finding on neuroimaging through transfontanelle ultrasounds, and/or computerized tomography, and/ or magnetic resonance imaging with identification of structural brain abnormalities, (i.e. disorders altering normal brain morphology).^2,3, 20^

### Later neurodevelopmental findings

Defined as abnormal based on the presence of any of the following beyond 6 months of life: (1) Severe developmental delay with a Bayley-III Score^21^ under -2SD (less than 70) in any functional domain (cognitive, language or motor) as previously described^4,5^; (2) Abnormal time to achievement of developmental stages through the Hammersmith Infant Neurological Examination scheme (HINES), evaluating neurological exam, motor function and state of behavior^4,5^.

### Statistical Analysis

Chi-square tests were used to examine differences in categorical covariates between groups. When cell counts were less than 5 infants for any group, Fisher-exact tests were performed. Positive and negative predictive values were calculated for below normal development based on clinical outcomes. Associations between gestational age at infection and clinical outcomes including microcephaly, structural image abnormalities, hearing or eye abnormalities, development, or ZIKV laboratory results were explored by Pearson’s Chi-Square. Analyses were conducted using the statistical package R (R version 3.0.1, The R Foundation for Statistical Computing, www.r-project.org).

### Study Oversight

The study was approved by the institutional review boards of the Fundação Oswaldo Cruz (Fiocruz) and the University of California, Los Angeles. Parents or guardians provided written informed consent.

## RESULTS

Our longitudinal cohort was comprised of 244 pregnant women with confirmed ZIKV infection during pregnancy, of whom 223 (91·4%) had live births. Of these, 216 infants had clinical follow-up beyond birth. In the early stages of the ZIKV epidemic in Rio de Janeiro between September 2015 to February 2016, ZIKV PCR and IgM detection assays were considered investigational and not diagnostic. For this reason, there was a delay in the collection of infant specimens while IRB approval for infant phlebotomy and urine collection was pending. As a result, from the original cohort of 216 infants, 130 children (60%) had blood and or urine specimens obtained for ZIKV detection. The present report focuses on these 130 infants with clinical follow-up with laboratory diagnostic evaluations. Table 1 reports clinical characteristics and timing of maternal infection for all 216 children in the cohort, with results stratified by those who received diagnostic testing and those who did not. As seen in the table both groups were comparable, with the exception that untested children tended to have lower neurodevelopmental scores in the second to third years of life.

**Table 1:** Frequency of abnormal findings among *in utero* ZIKV-exposed infants according to ZIKV postnatal testing status

|  | All infants<br>N=216 | % | Tested infants<br>N=130 | % | Untested<br>infants<br>N=86 | % | p-value** |
| --- | --- | --- | --- | --- | --- | --- | --- |
| Infants with any abnormal findings | 78/216 | 36.1 | 45/130 | 34.6 | 33/86 | 38.4 | p>0.05 |
| Brain Imaging with structural abnormalities | 14/140 | 10.0 | 14/113 | 12.4 | 0/27 | 0 | p>0.05 |
| Complete eye exam (Abnormal) | 9/137 | 6.6 | 6/109 | 5.5 | 3/28 | 10.7 | p>0.05 |
| Hearing Assessment (Abnormal) | 13/114 | 11.4 | 11/91 | 12.1 | 2/23 | 8.7 | p>0.05 |
| Small for gestational age | 10/216 | 4.6 | 10/130 | 7.7 | 0/86 | 0 | p=0.05 |
| Below average neurodevelopment* | 62/216 | 28.7 | 17/129 | 13.2 | 45/87 | 51.7 | p <0.00001 |
| 1st Trimester maternal infection | 54/216 | 25.0 | 40/130 | 30.7 | 14/86 | 16.3 | p=0.01 |
| 2nd Trimester maternal infection | 109/216 | 50.5 | 61/130 | 46.9 | 48/86 | 55.8 | p>0.05 |
| 3rd Trimester maternal infection | 53/216 | 24.5 | 29/130 | 22.3 | 24/86 | 27.9 | p>0.05 |
| Microcephaly | 8/216 | 3.7 | 8/130 | 6.1 | 0/86 | 0 | p>0.05 |

\*Assessed through Bayley-III or HINES
\*\* Chi-square (Fisher's Exact t-test used when cell values equal zero)

In total, 407 ZIKV diagnostic assays were run in specimens obtained from 130 exposed children. These included PCR assays in 161 serum and 85 urine samples, and 161 serum ZIKV IgM assays, as shown in Figure 1. In total, 84 of 130 children (65%) had positive laboratory test results for ZIKV as seen in Table 2. The age at the time of performance of the first Zika diagnostic laboratory test ranged from birth to 148 days. Within the group of children tested in the first 90 days of life (n=94), 70% (N=66) tested positive by at least one detection method (39% positive by blood PCR, 48% by urine PCR, 39% by IgM). For 78 children who were tested beyond 90 days of age, including repeat testing, 33% were positive by any laboratory detection method, demonstrating that sensitivity of diagnostic testing for ZIKV dropped considerably beyond 3 months of age. This observation was mainly due to the fact that the sensitivity of serum detection assays (IgM and PCR) in identifying Zika infection declined after 3 months of age (from 39% to 15% for serum PCR and from 39% to 20% for IgM). Although fewer children were tested by urine PCR (73 versus 109 for serum PCR), urine was the most frequently positive detection method within any age group (49%), followed by Zika IgM (positivity rate of 37%) and Zika PCR of serum (35%). (Table 2).

**Table 2:** ZIKV Vertical Transmission by age and assay type

|  | No. children | Positive | % |
| --- | --- | --- | --- |
| Tested within the first 3 months of age | 94 (72%) | 66 | 70% |
| PCR Serum | 76 (81%) | 30 | 39% |
| IgM | 75 (80%) | 29 | 39% |
| PCR Urine | 54 (57%) | 26 | 48% |
| First tested after 3 months of age* | 36 (28%) | 18 | 50% |
| PCR Serum | 33 (92%) | 7 | 21% |
| IgM | 36 (100%) | 7 | 19% |
| PCR Urine | 19 (53%) | 8 | 42% |
| Tested after 3 months of age | 78 (60%) | 26 | 33% |
| PCR Serum | 62 (79%) | 9 | 15% |
| IgM | 65 (83%) | 13 | 20% |
| PCR Urine | 23 (29%) | 10 | 43% |
| All time points | 130 (100%) | 84 | 65% |
| PCR Serum | 109 (84%) | 38 | 35% |
| IgM | 112 (86%) | 41 | 37% |
| PCR Urine | 73 (56%) | 36 | 49% |

\*94 children had their first laboratory test for ZIKV infection within 90 days of age; 36 children had their first assay performed after 90 days of age. Some children had one laboratory assay performed prior to 90 days of age and a subsequent laboratory assay performed post 90 days of age. Any children in this situation would be counted in the table as tested prior to 90 days of age. If there was repeat testing after 90 days of age children are also counted in that category for that specific assay, i.e., the number of assays does not reflect the number of children, as some children had more than one assay.

**Figure 1:**
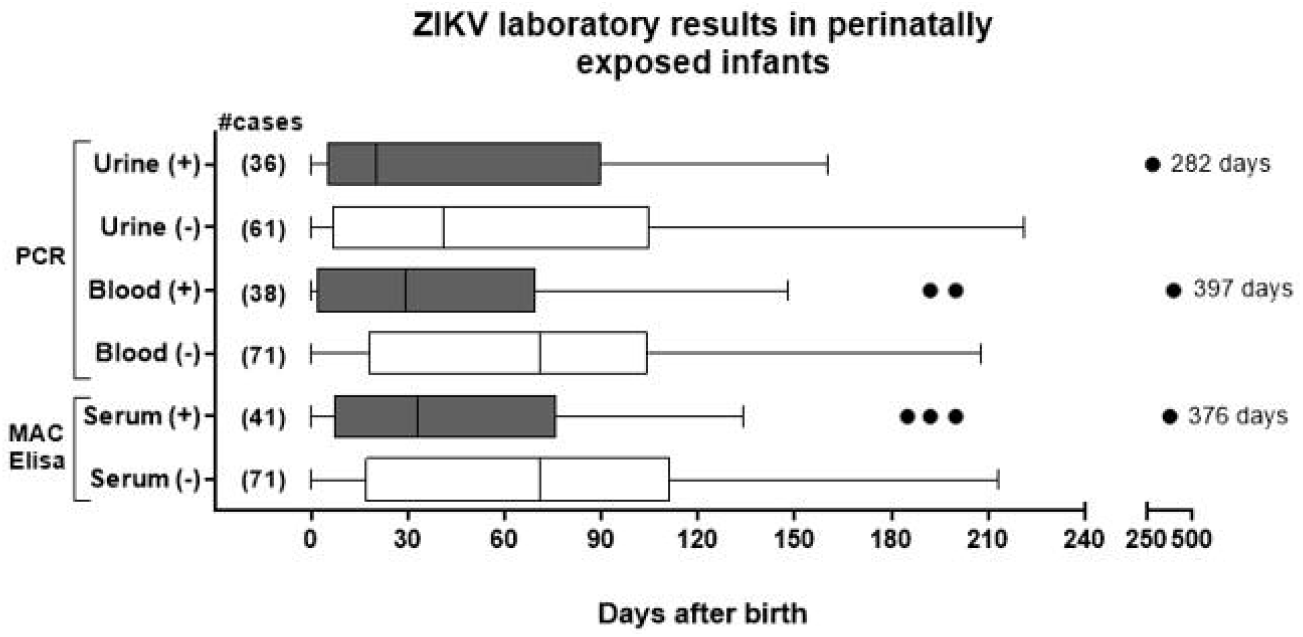
The figure reflects the number of assays performed in each assay category in specimens collected from 130 children. There were 161 serum specimens run for ZIKV PCR and ZIKV IgM and 85 urine specimens run for ZIKV PCR. The horizontal line in the box represents the median, the box the interquartile range (25%-75% of the data), and the whiskers the 95% confidence interval. The figure represents all the assays performed, not the number of children. Children can be represented more than once.

A positive ZIKV laboratory result in antenatally exposed children was most frequently seen in the first 3 months of life (Table 2 and Figure 1). Nevertheless, there were rare outliers within each assay category who tested positive beyond 200 days of age. Three children excreted the virus in urine at ages of 6·7, 8 and 9·4 months of age; none have developed abnormal clinical findings as of the time of this publication (3 years of age). However, two children with positive serum ZIKV PCR at ages 6 and 13 months were found to have abnormal funduscopic and hearing exams, respectively.

The frequency of specimen collection is depicted in Table 3. The majority of children were tested only once in each assay category, although blood specimens were obtained more than once in over 1/3 of children. Only one patient tested positive twice on a later time point by the same diagnostic assay (Zika IgM). Among children who had repeated testing at later time points in the first year of life or beyond, the overwhelming trend was for patients to remain negative or switch from positive to negative results (Table 3). Nevertheless, 2 children went from negative to positive results by serum assays and 3 children had positive results in urine by PCR after 2 previous negative assays.

**Table 3:** Frequency of Zika testing (n=407 in 130 children)

|  | No. of children | % | No. NEG every time | No. POS every time | No. changing from POS to NEG | No. changing from NEG to POS |
| --- | --- | --- | --- | --- | --- | --- |
| <u>Serum Zika RT-PCR*</u> | 109 |  |  |  |  |  |
| Once | 67 | 61.5% | 47 | 20 |  |  |
| 2 time points | 34 | 31.2% | 21 | 0 | 11 | 2 |
| 3 time points | 6 | 5.5% | 3 | 0 | 3 | 0 |
| 4 time points | 2 | 1.8% | 0 | 0 | 1 | 1 |
| <u>Zika IgM Mac Elisa**</u> | 112 |  |  |  |  |  |
| Once | 72 | 64.3% | 52 | 20 |  |  |
| 2 time points | 32 | 28.6% | 17 | 1 | 12 | 2 |
| 3 time points | 7 | 6.3% | 2 | 0 | 2 | 3 |
| 4 time points | 1 | 0.9% | 0 | 0 | 0 | 1 |
| <u>Urine Zika RT-PCR***</u> | 73 |  |  |  |  |  |
| Once | 64 | 87.7% | 34 | 30 |  |  |
| 2 time points | 6 | 8.2% | 3 | 0 | 3 | 0 |
| 3 time points | 3 | 4.1% | 0 | 0 | 0 | 3 |

\*Number of serum PCR tests (n=161)
\*\* Number of Zika IgM Mac Elisa\*\* (n=161)
\*\*\* Number of urine Zika RT-PCR\*\*\* (n=85)

Supplemental Table 1 depicts the different diagnostic assay combinations and concordance between different assays before or after 90 days of life. The majority of children (82·3%) had 2 or 3 concurrent diagnostic assays at their first visit. Over all time points, IgM results were concordant with serum and/or urine PCR results 52·3% of the time, with a sensitivity of 65·5% and a specificity of 39·6% (Supplemental Table 1). Concordance up to 90 days of age was 52% with a sensitivity of 61·5% and a specificity of 37·1%. Beyond age 3 months concordance for IgM and PCR assays was 39%, sensitivity 31·5% and specificity 35·7%. Concordance between IgM and serum PCR results was higher, 61% for all assays performed, while concordance between serum and urine PCR results was 55% at all time points.

Among the 130 children who were tested for ZIKV (Table 4), 113 (86%) had brain imaging, with 13% (n=14) demonstrating structural brain abnormalities (8 cases of microcephaly and 6 others with severe brain abnormalities). Complete eye exams were performed in 109 children in this cohort (83·8%); 6 children (5·5%) had abnormal eye findings. Ninety-one children (70%) had hearing assessments with 11 abnormal results (12·1%). Twenty-five children were noted to have grossly abnormal neurologic exams in the first six months of life (19%). Ten children (7·7 %) were born small for gestational age reflecting *in utero* growth restriction. Neurodevelopmental evaluations performed after one year of age demonstrated that 17 of 129 children (13·2%) had severe developmental delay, 14 scored below 2 SD in one or more Bayley-III domains and 3 were abnormal by the HINES assessment. When all parameters were combined, 45 of 130 children (35%) had an abnormal finding in one or more of these clinical or neurodevelopmental assessments.

**Table 4:** Association between ZIKV laboratory results, clinical findings and gestational age at infection

|  | All infants | Any Positive ZIKV-Lab result (N=84) |  | Any Negative ZIKV-Lab result (N=46) |  | p value |
| --- | --- | --- | --- | --- | --- | --- |
|  |  | n | % | n | % |  |
| All infants | 130 | 84 | 65% | 46 | 35% | 0.98 |
| Infants with abnormal findings* | 45 | 29 | 64% | 16 | 36% |  |
| Infants with no abnormal findings* | 85 | 55 | 65% | 30 | 35% |  |
| Brain Imaging | 113 | 74 | 65% | 39 | 35% | 0.37 |
| Structural abnormalities | 14 | 11 | 79% | 3 | 21% |  |
| Fundoscopy | 109 | 68 | 62% | 41 | 38% | 0.41 |
| Abnormal | 6 | 5 | 83% | 1 | 17% |  |
| Hearing Assessment | 91 | 60 | 66% | 31 | 34% | 0.09 |
| Abnormal | 11 | 10 | 91% | 1 | 9% |  |
| Neurologic exam | 130 | 84 | 65% | 46 | 35% | 0.94 |
| Abnormal | 25 | 16 | 64% | 9 | 36% |  |
| Gestational age assessment | 130 | 84 | 65% | 46 | 35% | 0.74 |
| Small for gestational age | 10 | 6 | 60% | 4 | 4% |  |
| Neurodevelopment | 129 | 83 | 64% | 46 | 36% | 0.76 |
| Abnormal Bayley-III or HINES | 17 | 12 | 71% | 5 | 29% |  |
| Gestational Trimester of Infection | 130 | 84 | 65% | 46 | 35% | 0.04 |
| 1st | 40 | 31 | 78% | 9 | 23% |  |
| 2nd | 61 | 39 | 64% | 22 | 36% |  |
| 3rd | 29 | 14 | 48% | 15 | 52% |  |

\* Includes abnormal hearing, eye, gestational age size assessment, neurodevelopment, neurological exam, or structural brain abnormality Pearson's Chi Square with Yates' continuity correction was used unless expected cell count was less than 5, then Fisher's exact test was used.

No statistical associations were identified between abnormal infant findings and positive ZIKV assay results, except for trimester of maternal infection. Infants born to mothers who contracted infection in the first trimester of pregnancy were more likely to have positive ZIKV PCR or IgM results (78%) as compared to those infected in the second (64%) or third trimesters (48%), p = 0·04 (Table 4). In addition, infants who were diagnosed by IgM in the first 90 days of life tended to have mothers who were infected earlier in gestation (median 14 weeks) than those diagnosed by IgM after 90 days of life (median 22 weeks), p < 0·01, as seen in Figure 2. The median week of infection during gestation did not differ between infants diagnosed by PCR in blood or urine before or after 90 days of age, (median 18 weeks for both groups), p =0·8470 Although no statistical association was identified between clinical abnormalities and a positive ZIKV test result (potentially because of the relatively small numbers in each category), we saw a tendency for specific abnormalities to cluster among children with positive results. Five of 6 infants with abnormal eye exams (83%), 11 of 14 with structural brain abnormalities (92%), and 10 of 11 with hearing deficits (91%) had a positive diagnostic assay for ZIKV infection. However, this clustering was not as clearly noted in children with other clinical findings such as developmental delay (12 of 17, or 71%), abnormal neurologic exams (16 of 25, 64%), or who were small for gestational age (6 of 10, 60%). None of these findings however achieved statistical significance as noted in Table 3. Overall 35% of the 130 children tested for ZIKV infection had at least one abnormal finding as seen in Table 3 which means the majority of the cohort (65%) had a normal outcome. Any positive laboratory test was found in 29 of 45 children (64%) with abnormal findings. Conversely, 55 of 85 children with no abnormalities (65%) also tested positive for ZIKV, demonstrating that a positive ZIKV test result was found in 2/3 of children with either normal or abnormal clinical findings.

**Figure 2:**
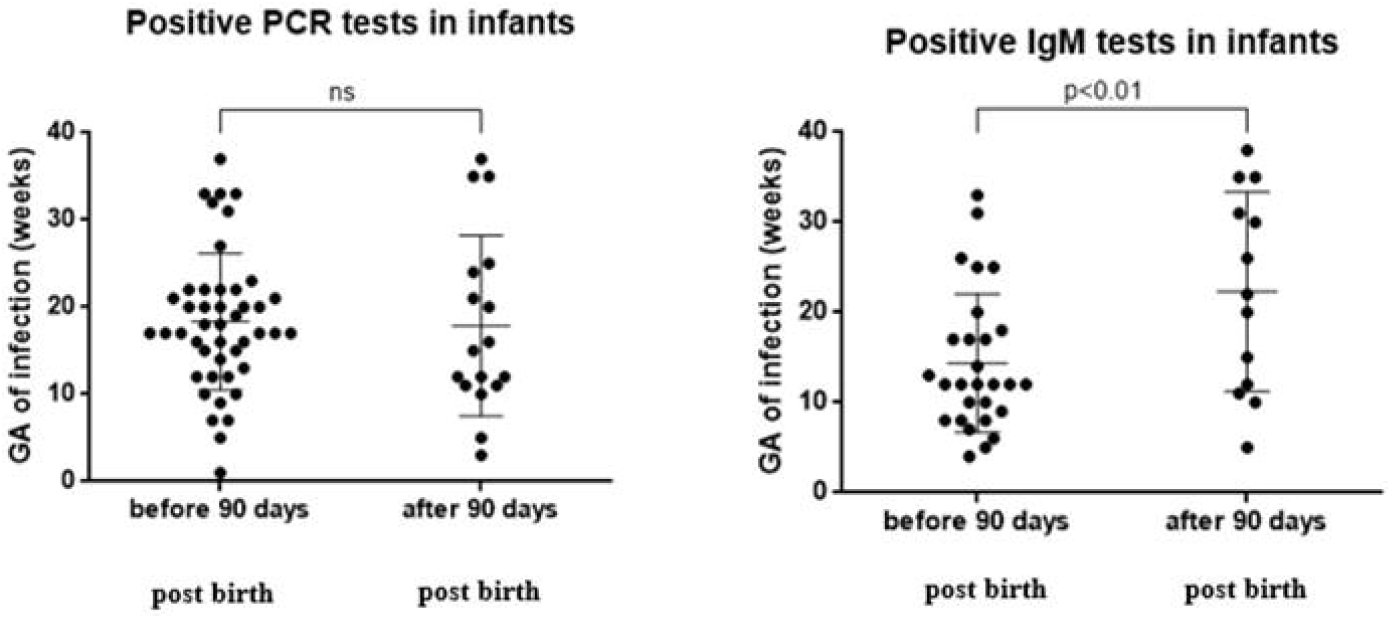
Infant positive PCR and IgM results by infant age and gestational age of infection

## DISCUSSION

Laboratory confirmation of ZIKV infection is challenging due to the short window of viremia and viruria enabling PCR detection, and also because of the serologic cross-reactivity between Zika and dengue viruses. In prenatally exposed children, a laboratory diagnosis of ZIKV infection is even more challenging. The duration of ZIKV viremia and viruria in congenitally infected children is unknown, and it is unclear if infants infected very early during intrauterine life have detectable virus at birth, as the duration of viral shedding from intrauterine infection has not been described. Additionally, it is unclear if viral presence in blood and/ or urine in congenitally infected children is constant or intermittent. It is unknown whether all children infected with ZIKV *in utero* form adequate antibody responses easily detectable by IgM assays. For example, experience from other congenital infections (such as rubella and CMV) tells us that IgM assays are suboptimal for diagnosis of viruses that affect T and B cell function. Nevertheless, IgM does not cross the placenta, so we therefore surmise that positive serologic results would reflect an infant’s prior exposure to the virus. In the present cohort, cross reactivity with dengue virus would be unlikely, as by the time infant specimens were obtained in Rio de Janeiro (2105-2017) there was absent to minimal circulation of dengue viruses.

The prospective nature of the cohort, with infants followed from the time of maternal infection, through birth, and onwards allows us a unique opportunity for evaluation of laboratory confirmed ZIKV congenital infection rates. Sixty-five percent of children in our cohort had laboratory evidence of ZIKV infection, including a large proportion of children who were infected in the first trimester of pregnancy (78%). Interestingly, having a positive ZIKV laboratory result did not necessarily correlate with infant outcomes; this could be due to limitations of the present sample size or a real phenomenon. What we can conclude from our study is that ZIKV testing of infants does not necessarily correlate with clinical findings, particularly in asymptomatic children. Anecdotally, in our cohort and that of other Zika cohorts in Brazil, there were symptomatic children tested in the first 48 hours of life who did not have detectable virus in either blood or urine. Potentially these children were infected so early during pregnancy that viral infection is gone by the time of birth and only the sequelae of infection is present. This is noted in other congenital infections such as congenital varicella syndrome.^22^ If infection is later in pregnancy, viral shedding may be more frequently seen, but the teratogenic sequelae will not be as obvious as the development of the neurologic system in the first 12 weeks of gestational age is past. So viral detection may not clearly correlate with the presence of clinical findings over time. This phenomenon is also seen in some children with congenital CMV.^23^ Children who acquire rubella after 20 weeks of gestation also do not have findings of congenital rubella syndrome.^24^ The high rate of positive laboratory results demonstrating infant ZIKV infection cannot be attributable to a high number of symptomatic children in the cohort. Eighty-five of 130 children in the study (65%) did not have any abnormal clinical manifestations at their last medical visit and did not have below average neurodevelopmental evaluations between ages 2 to 3 years. An equal proportion of children with both normal and abnormal findings tested positive (64 and 65% respectively). We conclude that ZIKV has a very high *in utero* transmission rate but laboratory confirmed ZIKV infection in an infant did not equate to the presence of severe congenital abnormalities in our cohort.

The transmission rate reported in our study is considerably higher than the transmission rate of 26% reported by colleagues in the French Guiana Zika infant cohort^25^. In that study, ZIKV-exposed children were clinically evaluated and tested for ZIKV in the first 7 days of life. Because the degree of intermittent viral shedding in the blood and urine in ZIKV infected infants has not been well characterized, testing for ZIKV over a longer period of time during infancy likely enhances detection of positive results. One study of pregnant women returning from epidemic areas to New York evaluated infant diagnosis around the time of birth, however the majority of maternal cases were suspected rather than PCR confirmed infections, with a 7% vertical transmission rate at birth for ZIKV infection reported^26^. Differences in study design (retrospective versus prospective, follow-up time, sampling, study population and definitions of infection, i.e., confirmed versus suspected) make it difficult to compare results across different studies.

Our specimens were collected and patients were recruited during the ZIKV epidemic, before there were any guidelines for ZIKV diagnostic evaluation of infants, including current CDC guidelines^27^. One could argue that the large number of positives could result from postnatal exposure to ZIKV. However, we should consider that all children in the present study had PCR-confirmed ZIKV *in utero* exposure, so they were all at high risk of contracting the virus. This differs significantly from scenarios in which the maternal diagnosis of Zika infection is unconfirmed and infant exposure status is unknown. Because ZIKV diagnostics in infants was considered investigational at the time, we did not collect specimens from infants born early on in the epidemic, which is when ZIKV was still circulating, which is a study limitation, as collection of early specimens in all children would have been ideal. In May 2016 we saw a dramatic decline in ZIKV circulation in Rio de Janeiro, which coincided with a Chikungunya outbreak^28-29^. For this reason, it is unlikely that the tested group of infants had a high chance of acquiring postnatal ZIKV infection, as by the time most of them were born, the virus was no longer circulating in Rio de Janeiro. We nevertheless did observe a much higher frequency of positive results in the first 3 months of life, reflecting a short window for detection of congenitally acquired ZIKV infection, including a short period of ZIKV IgM positivity, a finding that has been reported in adults following ZIKV infection^30^.

One of our study limitations is that we did not perform sequential testing of all infants at regular time points. We were able to perform PCR immediately following specimen collection for most infant serum and urine specimens which contributes to a greater sensitivity of the assay, as PCR identification of ZIKV tends to greatly diminish with freezing and thawing of specimens^31^. Nevertheless, because sequential, methodical testing at regular time points was not possible for many children, we cannot ascertain that the ones with negative results were true negatives. We certainly could have missed the window of positivity for a number of children who tested PCR negative for ZIKV. It is also unclear if all children exposed to ZIKV are capable of developing a robust IgM response. We know that in other congenital infections such as rubella and CMV, there can be a significant delay in the development of IgM antibodies^23-24^. In this sense, we believe the 65% ZIKV transmission rate is likely an underestimate.

We found a statistically significant association between earlier maternal infection in pregnancy and ZIKV infant infection, which demonstrates that transmission events are more likely when women are infected earlier in pregnancy. Nevertheless close to 50% of women infected in the third trimester also had infants with positive results, which underscores that ZIKV is highly transmissible throughout the entire gestational period. We also observed that women infected earlier in pregnancy tended to have infants with positive IgM results earlier in life. Conversely, women infected later in pregnancy tended to have infants whose IgM results were positive beyond 90 days of life. One caveat is that all women in our cohort were symptomatic. In a prior analysis we did not find any associations between the magnitude of maternal symptoms and infant clinical findings^32^. In addition, congenital Zika syndrome has been described in children of asymptomatic women^9^. Potentially women with symptomatic ZIKV infection could have children with more clinical findings, however this hypothesis has not yet been confirmed. Future research to determine factors related to transplacental transmission of the virus over gestation is needed.

As is the case with most congenitally acquired viral infections, such as CMV, rubella, or HIV, PCR of urine and serum (combined) was superior in diagnosing ZIKV infection as compared to serology alone. Urine PCR appeared to be most sensitive yielding the highest proportion of positive results^33^. As urine specimens were collected in 56% of the children in the cohort, higher rates of vertical transmission might have been observed if more urine specimens were obtained. IgM and serum PCR tended to provide similar rates of positivity. Nevertheless, because we did not have sequential, simultaneous testing of specimens at regular intervals by all assays, comparison between types of specimens and frequency of positive and negative results should be interpreted with caution.

Interestingly, a small number of children had positive PCR results after 200 days of life. One child developed positive IgM results during the same time period. Although we cannot rule out postnatal infection, because ZIKV stopped circulating in Rio de Janeiro by that time, postnatal infection seems unlikely. ZIKV could be shed intermittently in the blood or urine, similarly to congenital CMV or congenital rubella^23-24^. These children had negative test results before, which would suggest intermittent shedding. A delayed IgM response could be present in congenitally acquired ZIKV infection, explaining late positive IgM results. Intermittent and long-term shedding of ZIKV in the urine is very concerning for the presence of viral reservoirs with low-level replication that persist in some neonates.

A negative laboratory test result for ZIKV infection early in life will not rule out congenitally acquired infection because in many cases the narrow window period for testing may be missed. A positive result, however, particularly in our setting where women had PCR confirmed infection is helpful in ascertaining infection versus exposure. Current CDC testing guidelines recommend testing of infants as early as possible, preferably within the first few days after birth^23^, although according to our results testing specimens within the first few weeks up to 3 months of age might still be useful. Distinguishing between congenital, perinatal, and postnatal infection is difficult in infants living in endemic areas who are not tested soon after birth. Nevertheless, one has to take into account the epidemiologic surveillance data to ascertain if circulation of the virus is ongoing when interpreting results. There are no studies to date, to our knowledge, reporting data on sequential infant testing following PCR confirmed ZIKV antenatal exposure. Whether the pattern of viral shedding in antenatally infected infants is persistent or intermittent is unknown.

In summary, approximately 2/3 of children tested for ZIKV infection in our prospective cohort were positive, which means ZIKV *in utero* transmission is very frequent. Having a positive test result is not necessarily associated with a bad outcome, but there is an association with earlier maternal infection in pregnancy. Given the high transmission rate and the fact that a negative laboratory assay for ZIKV may not rule out infection, all infants with documented or potential antenatal ZIKV exposure should be followed long term. Intermittent viral shedding in the urine occurs in a small number of infants and suggests the presence of viral reservoirs. We have seen that a normal early infant assessment may not necessarily guarantee normal neurodevelopment or absence of sensory dysfunction^6^. As laboratory diagnostic testing does not seem to predict infant outcomes, close follow-up of all children with antenatal ZIKV exposure should be the norm.

## Data Availability

Data is available upon request

## ACKNOWLEDGMENTS

This study was supported by the Departamento de Ciência e Tecnologia (DECIT/25000.072811/2016-17) do Ministério da Saúde do Brasil (P.B., M.E.M.); Coordenação de Aperfeiçoamento de Pessoal de Nível Superior (CAPES/88887.116627/2016-01; CAPES 88881.130684/2016-01); Brazilian National Council for Scientific and Technological Development CNPq/441098/2016-9; CNPq307282/2017; CNPq440865/2016-6) (P.B., M.E.M., M.C.B); Fundação Carlos Chagas Filho de Amparo à Pesquisa do Estado do Rio de Janeiro (FAPERJ/ E_18/2015TXB (M.E.M.); FAPERJ/239224/E032018 CNE (M.E.M.); FAPERJ/ E_26/202.862/2018 CNE (P.B.); Fondation Christophe and Rodolph Mérieux; ZikAlliance 734548 (P.B.); the Thrasher Research Fund (20164370) (K.N.S., K.A.); the National Institute of Allergy and Infectious Diseases (NIAID) of the National Institutes of Health AI28697 (K.N.S.), AI1259534-01 (K.N.S., P.B., G.C.), AI140718-01 (K.N.S., P,B.), K08AI141728 (S.L.G), the National Eye Institute (NEI) of the National Institutes of Health AI129847-01 (K.N.S., I.T, P.B.) and the United Kingdom’s Department for International Development (M.E.M.); the Queenan Fellowship from the Foundation for the Society of Maternal-Fetal Medicine (S.L.G) and the ZikaPlan (Preparedness Latin American Network) (M.E.M.); Wellcome Trust & the UK’s Department for International Development (205377/Z/16/Z; https://wellcome.ac.uk/) and the European Union’s Horizon 2020 research and innovation program (https://ec.europa.eu/programmes/horizon2020/) under ZikaPLAN grant agreement No. 734584 (https://zikaplan.tghn.org/) (E.B.)

We thank the women who enrolled in this study and the Fiocruz Zika Field Team who rendered our work possible.

## AUTHOR CONTRIBUTIONS

P.B., Z.V., K.N.-S., C.R.G., and M.E.M. conceived and designed the study. P.B, K.N.-S, T.K., I.P.R., M.C.B., Z.V., L.D., L.C., D.C., M.P., S.P, A.Z., I.T., M.E.M., L.W., S.A., G.C., S.F., W.C., J.J. were responsible for data collection and accuracy checks of data. P.B, T.K., Z.V., K.N.-S, were responsible for data analysis. P.B, K.N.-S, T.K., Z.V., I.R., M.B., K.A., L.D., A.Z., I.T., J.P.P., S.L.G., J.D.C., L.G., M. R., E.B., M.E.M., G.C., J.J. were responsible for interpreting the data. P.B, Z.V., K.N.-S, T.K., C.R.G. drafted the manuscript. All authors critically revised the manuscript and gave final approval of the version to be published.

## COMPETING INTEREST STATEMENT

The authors declare no competing interests.

## Notes

### Competing Interest Statement

The authors have declared no competing interest.

## REFERENCES

1. Calvet GA, Filippis AMB, Mendonça MCL, et al. First detection of autochthonous Zika virus transmission in a HIV-infected patient in Rio de Janeiro, Brazil. J Clin Virol 2016; 74: 1–3.

2. Brasil P, Pereira JP Jr, Moreira ME, et al. Zika Virus Infection in Pregnant Women in Rio de Janeiro - Preliminary report. N Engl J Med Epub 2016 March 4.

3. Brasil P, Pereira JP Jr, Moreira ME, et al. Zika Virus Infection in Pregnant Women in Rio de Janeiro. N Engl J Med 2016; 375: 2321–34.

4. Lopes Moreira ME, Nielsen-Saines K, Brasil P, et al. Neurodevelopment in Infants Exposed to Zika Virus In Utero. N Engl J Med 2018; 379: 2377–9.

5. Nielsen-Saines K, Brasil P, Kerin T, et al. Delayed childhood neurodevelopment and neurosensory alterations in the second year of life in a prospective cohort of ZIKV-exposed children. Nature Medicine. 2019. DOI:10.1038/s41591-019-0496-1.

6. Einspieler C, Utsch F, Brasil P, et al. Association of Infants Exposed to Prenatal Zika Virus Infection With Their Clinical, Neurologic, and Developmental Status Evaluated via the General Movement Assessment Tool. JAMA Netw Open 2019; 2: e187235.

7. Adachi K, Nielsen-Saines K. Zika clinical updates: implications for pediatrics. Curr Opin Pediatr 2018; 30: 105–16.

8. Pereira JP Jr, Maykin MM, Vasconcelos Z, et al. The Role of Amniocentesis in the Diagnosis of Congenital Zika Syndrome. Clin Infect Dis 2019; published online Jan 8. DOI:10.1093/cid/ciz013.

9. Paz-Bailey G, Rosenberg ES, Doyle K, et al. Persistence of Zika Virus in Body Fluids — Final Report. New England Journal of Medicine. 2018; 379: 1234–43.

10. Sánchez-Montalvá A, Salvador F, Molina I. Persistence of Zika Virus in Body Fluids — Final Report. N Engl J Med 2019; 380: 198.

11. Braga JU, Bressan C, Dalvi APR, et al. Accuracy of Zika virus disease case definition during simultaneous Dengue and Chikungunya epidemics. PLoS One 2017; 12: e0179725.

12. Lanciotti RS, Kosoy OL, Laven JJ, et al. Genetic and serologic properties of Zika virus associated with an epidemic, Yap State, Micronesia, 2007. Emerg Infect Dis 2008; 14: 1232–9.

13. Granger D, Hilgart H, Misner L, et al. Serologic Testing for Zika Virus: Comparison of Three Zika Virus IgM-Screening Enzyme-Linked Immunosorbent Assays and Initial Laboratory Experiences. J Clin Microbiol 2017; 55: 2127–36.

14. Hagen M von der, von der Hagen M, Pivarcsi M, et al. Diagnostic approach to microcephaly in childhood: a two-center study and review of the literature. Dev Med Child Neurolog. 2014; 56: 732–41.

15. Villar J, Cheikh Ismail L, Victora CG, et al. International standards for newborn weight, length, and head circumference by gestational age and sex: the Newborn Cross-Sectional Study of the INTERGROWTH-21st Project. Lancet 2014; 384: 857–68.

16. Tsui I, Moreira MEL, Rossetto JD, et al. Eye Findings in Infants With Suspected or Confirmed Antenatal Zika Virus Exposure. Pediatrics. 2018; 142: e20181104.

17. Zin AA, Tsui I, Rossetto J, et al. Screening Criteria for Ophthalmic Manifestations of Congenital Zika Virus Infection. JAMA Pediatrics. 2017; 171: 847.

18. Freitas B de P, de Paula Freitas B, Zin A, et al. Anterior-Segment Ocular Findings and Microphthalmia in Congenital Zika Syndrome. Ophthalmology. 2017; 124: 1876–8.

19. Zin AA, Tsui I, Rossetto JD, et al. Visual function in infants with antenatal Zika virus exposure. J Assoc Ped Ophthal Strabismus. 2018; 22: 452–6.e1.

20. Pool KL, Adachi K, Romero T, et al. Association Between Neonatal Neuroimaging and Clinical Outcomes in Zika-Exposed Infants from Rio de Janeiro, Brazil. JAMA Network Open. 2019; 2: e198124.

21. Johnson S, Moore T, Marlow N. Using the Bayley-III to assess neurodevelopmental delay: which cut-off should be used? Pediatr Res. 2014; 75: 670–4.

22. Pastuszak AL, Levy M, Schick B, et al. Outcome after maternal varicella infection in the first 20 weeks of pregnancy. N Engl J Med. 1994; 330: 901–5.

23. Harrison GJ. Cytomegalovirus. In: Cherry JD, Harrison GJ, Kaplan SL, et al, ed. Feigin and Cherry’s Textbook of Pediatric Infectious Diseases. Elsevier Saunders, 2019: 1429.

24. Cherry JD, Baker A. Rubella virus. In: Cherry JD, Harrison GJ, Kaplan SL, et al, ed. Feigin and Cherry’s Textbook of Pediatric Infectious Diseases. Elsevier Saunders, 2019: 1601.

25. Pomar L, Vouga M, Lambert V, et al. Maternal-fetal transmission and adverse perinatal outcomes in pregnant women infected with Zika virus: prospective cohort study in French Guiana. BMJ. 2018; 363: k4431.

26. Conners EE, Lee EH, Thompson CN, et al. Zika Virus Infection Among Pregnant Women and Their Neonates in New York City, January 2016-June 2017. Obstet Gynecol. 2018; 132: 487–495.

27. Adebanjo T, Godfred-Cato S, Viens L, et al. Update: Interim Guidance for the Diagnosis, Evaluation, and Management of Infants with Possible Congenital Zika Virus Infection - United States, October 2017. MMWR Morb Mortal Wkly Rep 2017; 66: 1089–99.

28. Sharp TM, Fischer M, Muñoz-Jordán JL, et al. Dengue and Zika Virus Diagnostic Testing for Patients with a Clinically Compatible Illness and Risk for Infection with Both Viruses. MMWR Recomm Rep 2019; 68: 1–10.

29. Fuller TL, Calvet G, Estevam CG, et al. Behavioral, climatic, and environmental risk factors for Zika and Chikungunya virus infections in Rio de Janeiro, Brazil, 2015-16. PLOS ONE. 2017; 12: e0188002.

30. Theel ES, Hata DJ. Diagnostic Testing for Zika Virus: a Post-outbreak Update. J Clin Microbiol 2018; 56. DOI:10.1128/JCM.01972-17.

31. Tan SK, Sahoo MK, Milligan SB, Taylor N, Pinsky BA. Stability of Zika virus in urine: Specimen processing considerations and implications for the detection of RNA targets in urine. J Virol Methods 2017; 248: 66–70.

32. Halai UA, Nielsen-Saines K, Moreira ML, et al. Maternal Zika Virus Disease Severity, Virus Load, Prior Dengue Antibodies, and Their Relationship to Birth Outcomes. Clin Infect Dis. 2017; 65: 877–883.

33. Vasconcelos Z, Azevedo RC, Zin A, et al. ZIKV Diagnostics: Current Scenario and Future Directions. Biochemical Testing - Clinical correlation and Diagnosis. 2018; Last accessed 2/22/2020: https://www.intechopen.com/online-first/zikv-diagnostics-current-scenario-and-future-directions/

